# Optimization of Microbiological Laboratory Detection Strategy for Patients in A Designated Hospital Treating Novel Coronavirus Pneumonia in Anhui Province

**DOI:** 10.1101/2020.03.21.20039065

**Authors:** Wenjiao Chang, Yuru Shi, Yingjie Qi, Jiaxing Liu, Ting Liu, Zhaowu Chen, Wenjing Zhang, Mengmeng Wang, Dongfeng Liu, Ming Yin, Jing Xu, Yun Yang, Xiaowu Zhu, Jing Ge, Shu Zhu, Yong Gao, Xiaoling Ma

**Author notes:** Corresponding Author: Xiaoling Ma, The First Affiliated Hospital of USTC, Division of Life Sciences and Medicine, University of Science and Technology of China, Hefei, Anhui, 230000, China (., Yong Gao, The First Affiliated Hospital of USTC, Division of Life Sciences and Medicine, University of Science and Technology of China, Hefei, Anhui, 230000, China (.

## Abstract

Novel coronavirus pneumonia (NCP) is an emerging, highly contagious community acquired pneumonia (CAP) caused by severe acute respiratory syndrome coronavirus 2 (SARS-CoV-2). Highly efficient and accurate microbiological laboratory assay is essential to confirm the SARS-CoV-2 infection, rule out other pathogens that can cause CAP, and monitor secondary infections. Here, we enrolled and provide microbiological analysis for 129 suspected and 52 transferred confirmed NCP patients hospitalized in the First Affiliated Hospital of University of Science and Technology of China (USTC) from Jan 21 to Feb 29, 2020. By analyzing the dual swab samples (sputum and pharyngeal) from 129 suspected patients with realtime RT-PCR, we confirmed 33 SARS-CoV-2 infections, with two co-infection cases with adenovirus or rhinovirus. We also used multiplex PCR to detect 13 common respiratory tract pathogens in 96 non-NCP patients, and found that 30 patients (31.25%) were infected with at least one respiratory tract pathogen that may cause CAP. Further, we performed bacterial and fungal cultures as well as fungal serologic tests and found that there is no secondary bacterial/fungal infections in confirmed NCP patients. Our studies suggest that, during the epidemic of NCP in Anhui province, there was a certain proportion of infection and co-infection of other common pathogens of CAP, and the secondary bacterial and fungal infection is not detectable in NCP patients. In comparison with SARS-CoV-2 detection alone, this optimized strategy combining multiple pathogen detection for identification of NCP and other CAP patients as well as cultures and serologic tests for confirmed patients increases the diagnosis efficiency and facilitates the personalized medication.

## Introduction

In December 2019, a series of pneumonia cases of unknown origins were reported in Wuhan, Hubei Province, and a novel beta-coronavirus was subsequently identified from the respiratory specimens of the patients by High-throughput sequencing[1]. The virus was confirmed as the cause of the unexplained pneumonia and named as severe acute respiratory syndrome coronavirus 2 (SARS-CoV-2), which has resulted in a rapid outbreak throughout China and global spread, and posed a significant threat to public health[2-6]. According to a recent study, the transmission capacity of SARS-CoV-2 was higher than the severe acute respiratory syndrome coronavirus (SARS-cov)[7]. In the meantime, the infection by the virus could cause substantial critical conditions (14.8%∼15.74%), as well as significant mortality (1.4%∼14.6%)[8-10]. Therefore, early diagnosis, early isolation and early management are crucial to curb rapid spread of the virus and improve the prognosis of the patients[11].

Novel coronavirus pneumonia (NCP) is an emerging community acquired pneumonia (CAP). The clinical symptoms of NCP include fever, dry cough, muscle pain and so on[8]. Ground-glass opacity and bilateral patchy shadowing were the radiological features of the NCP patients[8, 12]. Whereas some pathogens, such as RSV, parainfluenza virus may share similar imaging features (Figure 1). SARS-CoV-2 nucleic acid test is recommended as the standard for diagnosis of the disease. However, this method is often affected by many factors such as sample collection and reagent quality. False negative results could occur, resulting in delayed or missed diagnosis[9]. In this study, both sputum and pharyngeal swab samples were collected, then mixed and detected for SARS-CoV-2 using two different real-time RT-PCR reagents, which significantly improved the positive rate of detection.

**Figure 1.**
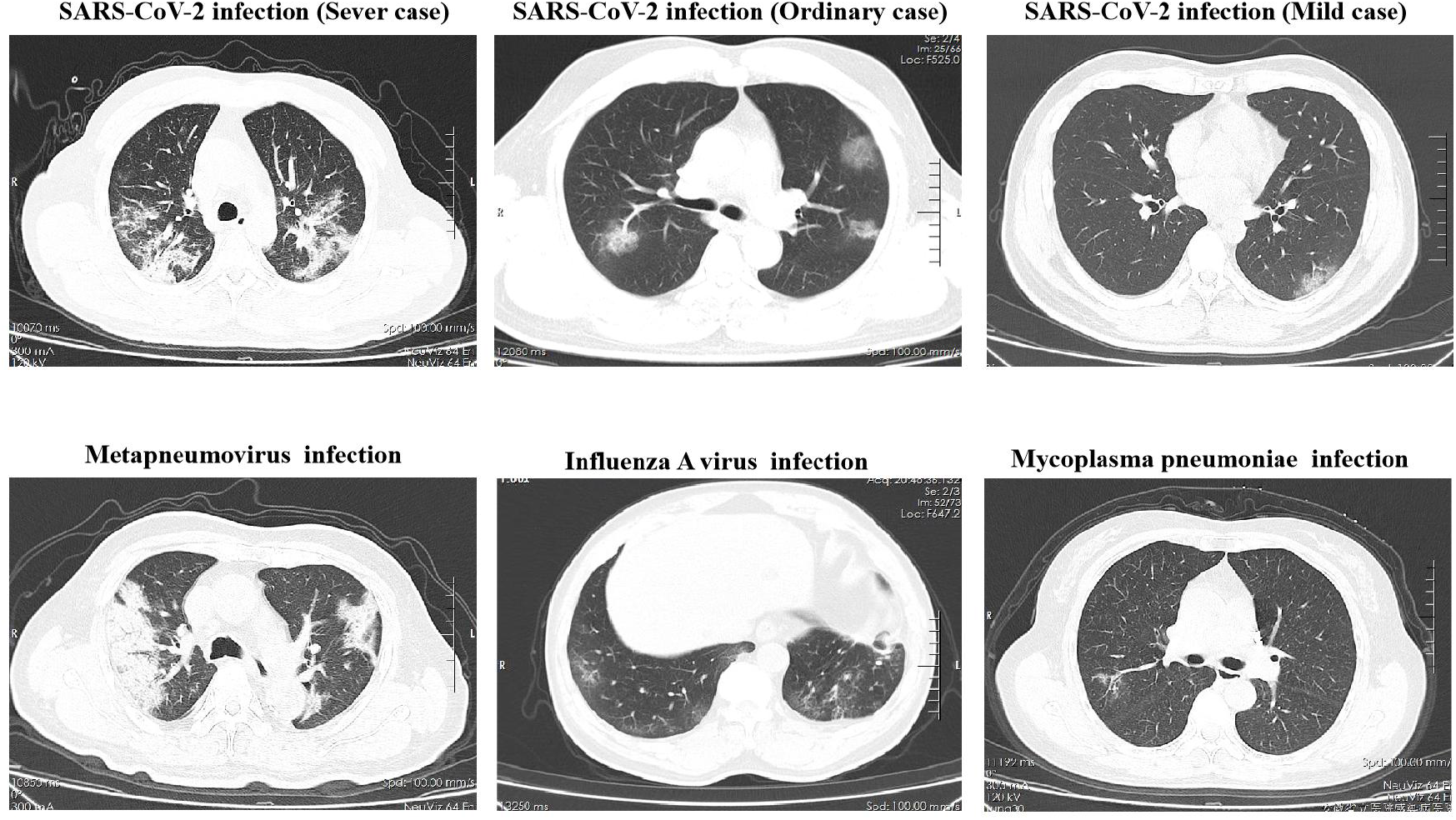
Chest computed tomographic Images of patients with COVID-19 and other pathogens.

The winter is the season of high incidence of CAP, SARS-CoV-2 nucleic acid detection can’t clearly suggest which pathogen is responsible for the SARS-CoV-2 negative patients, nor can it determine whether the positive patients have co-infection. In order to improve the etiological diagnosis, 13 different respiratory tract pathogens (RTPs) along with SARS-CoV-2 were simultaneously detected in suspected NCP patients.

According to the Chinese “New Coronavirus Pneumonia Prevention and Control Program”[13], a second nucleic acid test is performed after 24 hours, and only when both nucleic acid test results are negative can the patient be discharged. The average hospitalization time of NCP patients was reported more than 14 days[14]. In order to monitor whether secondary nosocomial infection occurs during hospitalization, we used bacterial and fungal cultures and fungal serological tests to monitor secondary infection in hospitalized patients with fever, increased sputum and increased inflammatory factors.

In this study, we used different microbiological laboratory detection strategies for suspected NCP patients and hospitalized confirmed patients, which allowed rapid diagnosis of infection and co-infection with SARS-CoV-2 and other common pathogens of community-acquired pneumonia. The results of microbial laboratory detections can guide the clinical precise treatment and reduce the unnecessary use of antibiotics.

## Materials and Methods

### Patients

We recruited 181 patients with suspected SARS-CoV-2 infection from Jan 21 to Feb 29, 2020 at the First Affiliated Hospital of University of Science and Technology(USTC) of China in Hefei, China. This hospital is designated to treat NCP patients in Anhui Province. All of the suspected patients enrolled in this study met the criteria of the latest “New Coronavirus Pneumonia Prevention and Control Program”[13]. Written consents were obtained from all of the patients and the study was approved by the Ethics Review Committee of the First Affiliated Hospital of USTC.

### Microbial etiological diagnostic strategy for suspected NCP patients

#### 1. Detection of novel coronavirus nucleic acid

Both Sputum and pharyngeal swabs were collected from all suspected patients on the admission day, then mixed. Total RNA was extracted from specimens using an automatic nucleic acid extraction instrument (TANBead, China), SARS-CoV-2 was then detected by real-time RT-PCR using two different detection kits. One kit selected *ORF1ab* gene as the target sequence for detection (Beijing Genomics Institution, China), and the other one used *ORF1ab, E gene and N gene* as the target sequences (Bioperfectus technologies, China). In brief, the total RNA was added to the reaction reagent and dual fluorescence PCR (Applied Biosystems 7500 Real-Time PCR Systems, USA) was performed according to the manufacturer’s instructions. If the first SARS-CoV-2 real-time RT-PCR result is negative, the respiratory specimens will be collected again after 24 hours for another test.

#### 2. Detection of common pathogens in community acquired pneumonia using the SureX 13 respiratory pathogen multiplex kit

The SureX 13 respiratory pathogen multiplex kit (Ningbo Health Gene Technologies Ltd Ningbo, China) was used to detect 13 common pathogens of CAP, including influenza A virus (FluA), pandemic influenza A virus-2009 (09H1), seasonal H3N2 virus (H3), influenza B virus (FluB), respiratory syncytial virus (RSV), adenoviruses (AdVs), rhinovirus (HRV), bocavirus (HBoV), metapneumovirus (HMPV), parainfluenza virus (PIV, including PIV-1, 2, 3 and 4), coronavirus (CoV, including OC43, 229E, NL63 and HKU1), Chlamydia [Ch, including Chlamydia trachomatis (Ct) and Cp] and Mycoplasma pneumoniae. In brief, the extracted nucleic acid was mixed with primer pairs targeting the 13 tested RTPs and RT-PCR enzyme mix, and the PCR amplification was performed in Biosystems 7500 Real-Time PCR Systems, followed by capillary electrophoresis. PCR products were then separated by size on the Applied Biosystems 3500Dx Genetic Analyzer. The signals of the 15 labeled PCR products were finally measured by fluorescence and analyzed with GeneMapper 4.1 software (Termo Fisher Scientific, Waltham, MA, USA).

### Monitoring strategy for secondary infection in NCP patients during hospitalization

#### 1. Culture

If a patient had fever, aggravated cough, imaging changes or increased inflammatory factors in the course of treatment, sputum sample would be obtained for identification of possible causative bacteria or fungi. All specimens were plated onto sheep blood agar (SBA), chocolate agar, MacConkey agar and Sabouraud agar. The Sabouraud agar plates were incubated at 27 °C for 5 days and the other plates were held for 2 days at 35°C. Blood samples were collected and injected into aerobic and anaerobic blood culture bottle. The culture of aerobic and anaerobic organisms was performed in the BACT ALERT 3D (BioMerieux, France). When the blood culture system reported positive, the corresponding blood sample was then subcultured onto blood agar and sabouraud agar plates. Plates were held at 35 °C for 2 days and checked for colony formation.

#### 2. Fungal serologic tests

If fungal infection was suspected in a patient, the blood samples would be obtained for GM test (Dynamiker Biotechnology, Tianjin, China) and for detection of Aspergillus galactomannan antigen. In brief, the serum samples were added into wells precoated with galactomannan antibody and then incubated. The wells were washed to remove the unbound material, and then added the conjugate and incubated. Finally, the substrate solution was added into the wells for color development which was terminated by adding the stopping solution. The absorbance (optical density) of specimens and controls was determined with a spectrophotometer at 450 and 620/630 nm wavelength.

G test offers a diagnostic reference for invasive fungal diseases. To carry out the test, the pre-treated sera were added into the Main Reagent containing Factor G. The latter was activated by (1-3)-β-D-Glucan and converted from proclotting enzyme into clotting enzyme which hydrolyzed the substrate (Boc-leu-Gly-Arg-PNA) to release PNA. The absorbance was measured at 405nm kinetically, and the concentration of (1-3)-β-D-Glucan was interpreted according to a standard curve.

### Follow-up

Clinical data were obtained from patients’ medical records. The patients were followed up until Mar 8, 2020.

## Results

In total, 181 patients were enrolled in this study. Among them, 129 were suspected NCP patients and hospitalized for further diagnosis and treatment, and 52 patients were diagnosed as SARS-CoV-2 infection in other medical institutions and transferred to our hospital.

### Dual reagents screening with mixed sputum and pharyngeal swab samples improved the sensitivity of detection of SARS-CoV-2

Of the 129 suspected patients, 33 patients were confirmed of SARS-CoV-2 infection by using our dual specimen and dual reagents screening strategy through real-time RT-PCR assays. Of these 33 confirmed patients, 31 (93.93%) patients were positive in the first test. The 96 excluded patients were observed in hospital and followed up after discharge, and no additional NCP was found.

### Detection of pathogens other than SARS-CoV-2 in community acquired pneumonia

129 suspected patients were detected for 13 respiratory tract pathogens other than SARS-CoV-2. Of the 33 NCP patients, one patient was also infected with adenovirus and the other with rhinovirus. Of the 96 non-NCP patients, common CAP pathogens were detected in 30 patients. Among them, one single pathogen infection was detected in 28 patients, and two pathogens were found in 2 patients. The most common pathogens were *influenza A virus* (18.75%), *influenza B virus* (18.75%), *Mycoplasma pneumoniae* (18.75%) and *metapneumovirus* (15.63%), followed by several other pathogens (Figure 2 and 4).

**Figure 2.**
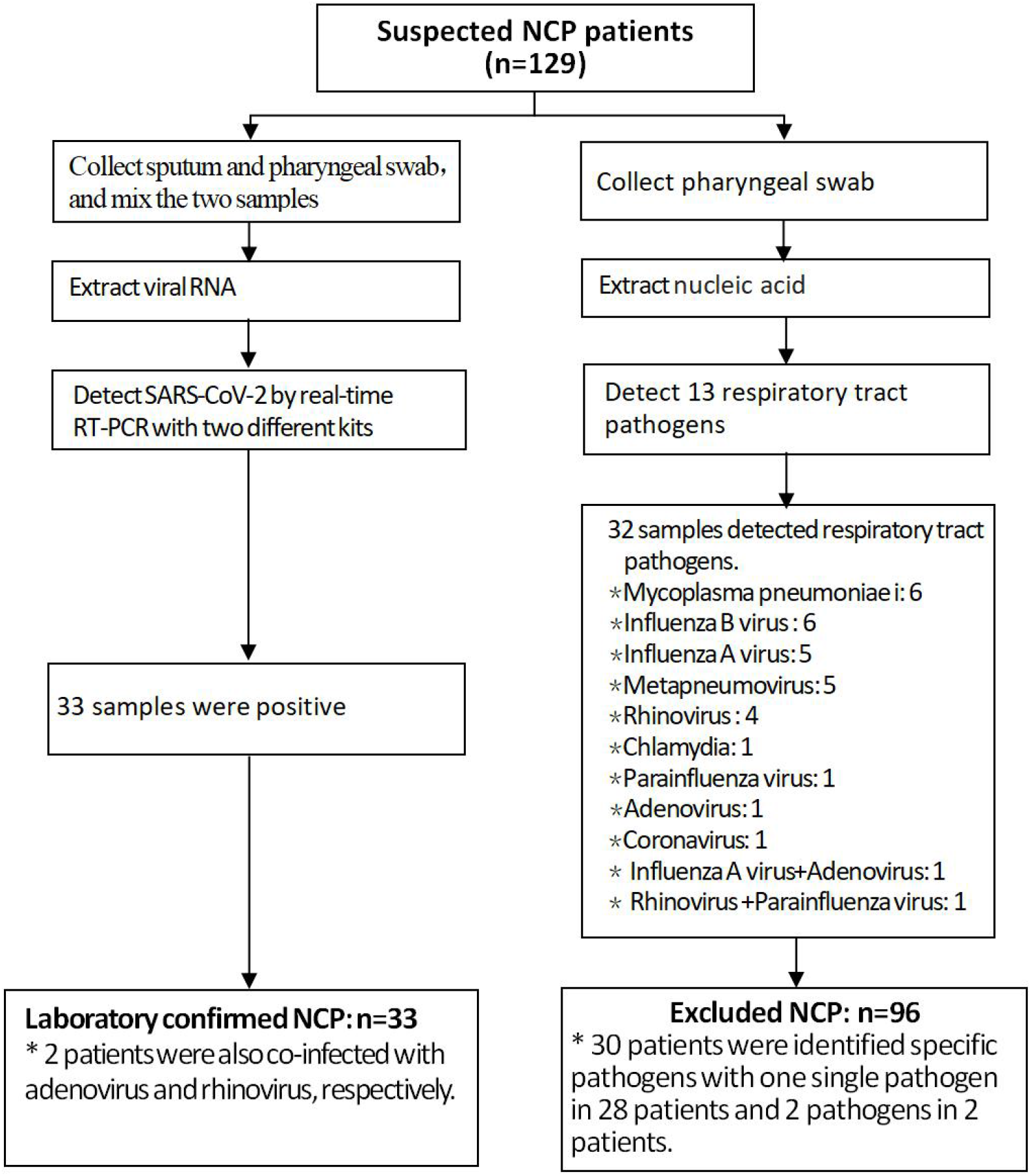
Flow chart for differential diagnosis of 129 suspected NCP patients

**Figure 3.**
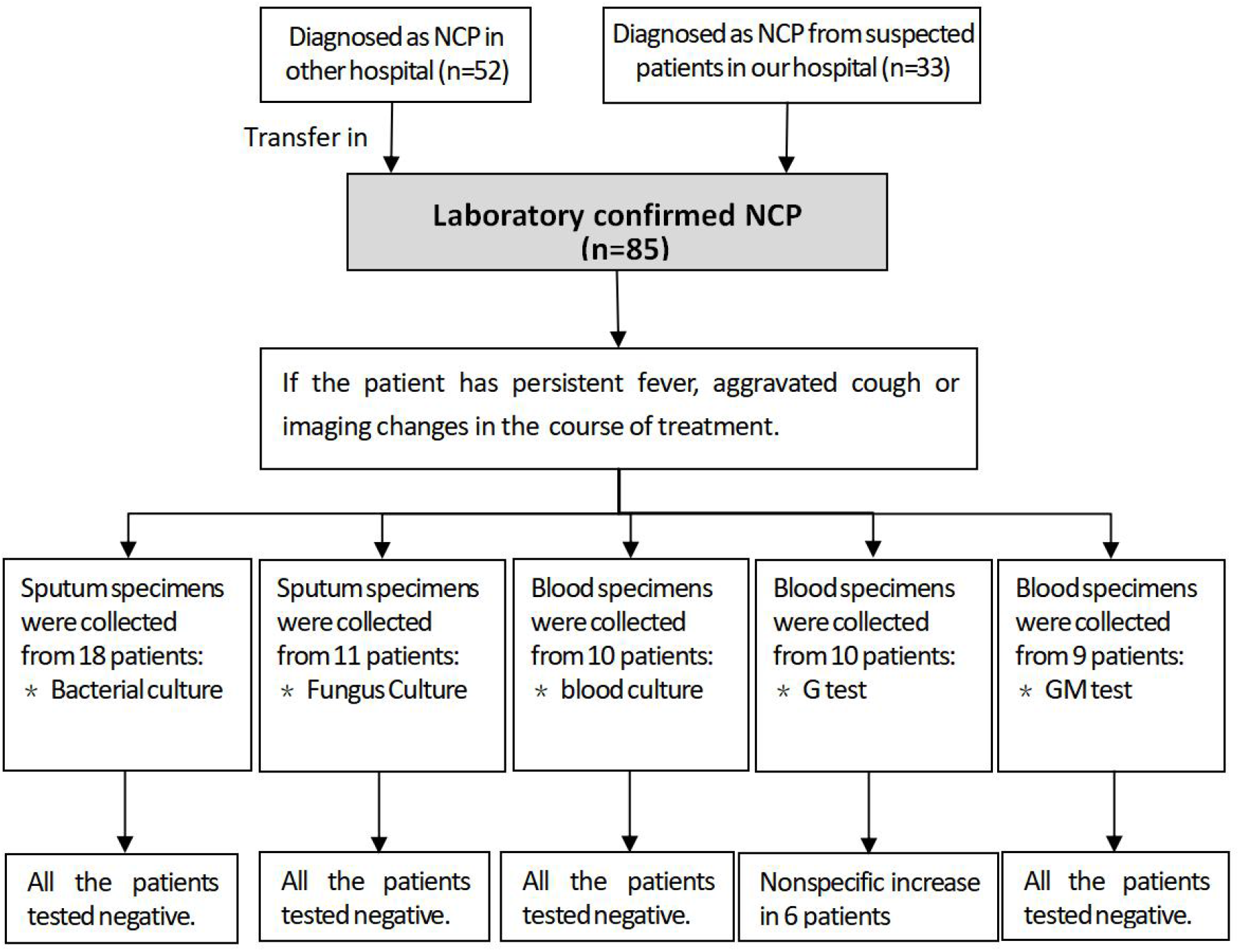
Flow chart of monitoring secondary infection during treatment in laboratory confirmed NCP patients

**Figure 4.**
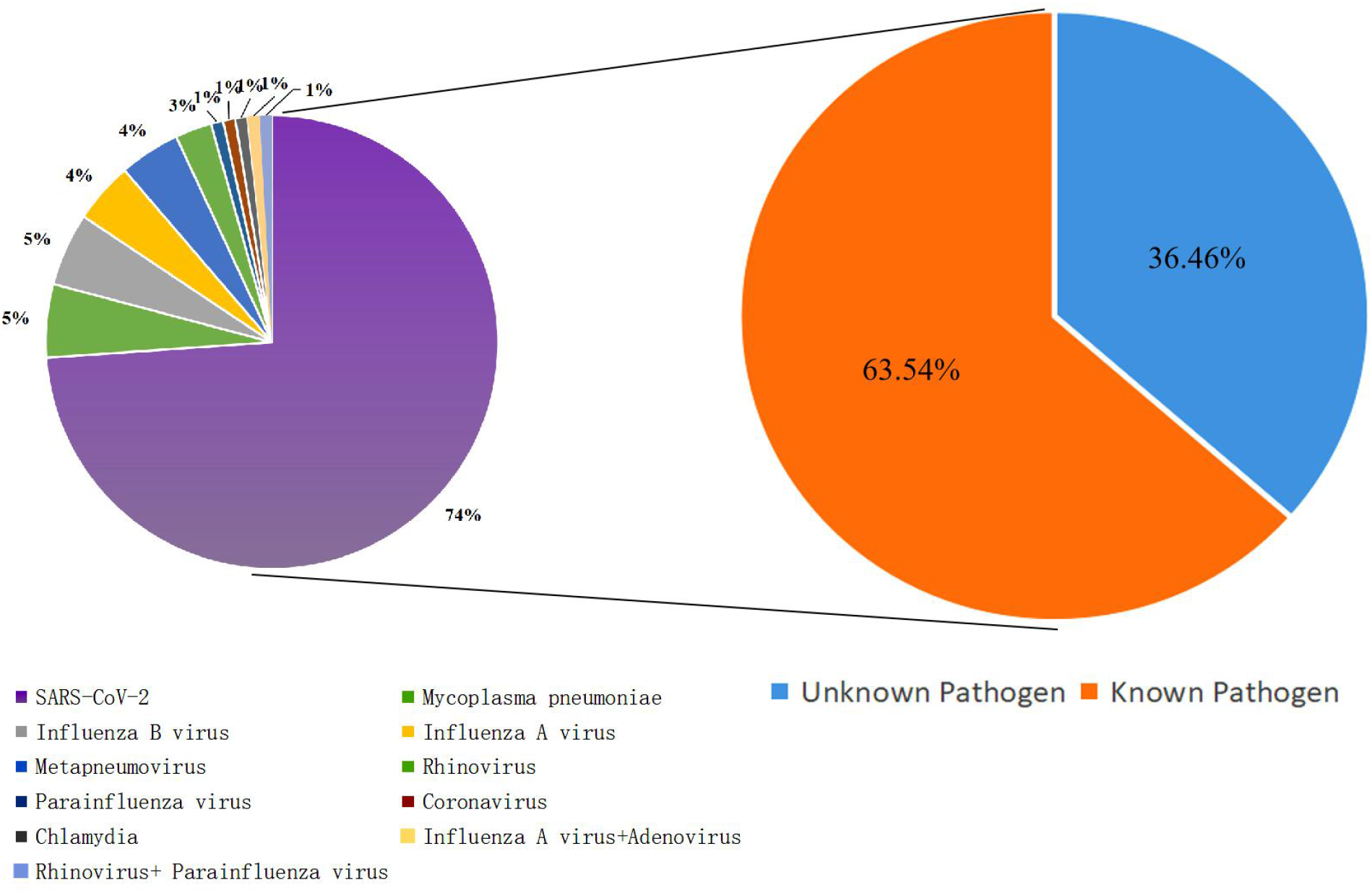
Distribution of pathogens in 181 hospitalized patients

### Bacterial and fungal cultures

Blood cultures for 10 NCP patients, bacterial and fungal cultures for sputum samples from 18 and 11 NCP patients, respectively, were performed. No bacteria or fungi were isolated from any of these tested samples (Figure 3).

### Fungal serologic tests

GM test was done in 9 NCP patients, and the results were all negative. However, G test showed a nonspecific elevation in 6 out of 10 tested NCP patients (Figure 3).

### Treatment and outcome of patients

NCP patients were treated based on the severity of the disease according to latest “New Coronavirus Pneumonia Prevention and Control Program”. Non-NCP patients with a definite microbial diagnosis were treated according to the international community acquired pneumonia guidelines. By Mar 8 of 2020, 84 (98.82%) NCP patients was discharged from hospital (including 29 patients with severe NCP) and 1 (1.17%) patient died from the disease. The average hospitalization time of NCP patients was 16.79days. The dead case was a 55-year-old man and was transferred to our hospital after confirmed as SARS-CoV-2 infection.

He was diagnosed with severe pneumonia, severe respiratory failure and cerebral apoplexy. One day after admission to our intensive care unit (ICU), the patient died from a sudden cardiac arrest. 91 (94.79%) non-NCP patients were discharged from hospital, 4 (4.17%) were transferred to other hospitals for further treatment, and 1 (1.04%) died.

## Discussion

The SARS-CoV-2 is a positive-sense, single-stranded RNA virus and is the seventh member of enveloped RNA coronavirus[4, 5]. The SARS-CoV, which caused an epidemic in 2002-2003, also belongs to this genus. Infection with these two viruses often leads to lower respiratory tract disease with poor clinical outcomes associated with old age and underlying health[15], while SARS-CoV-2 seems to be more transmissible than SARS-CoV[7]. The virus is transmitted mainly through respiratory droplets or close contact. A recent study revealed that the R0 of SARS-CoV-2 is 3.77, meaning that each infector could transmit the virus to another 3.77 people, which is higher than the R0 of SARS (2.0-3.0)[16]. As of Mar. 12^th^ of 2020, data from the World Health Organization (WHO) have shown that more than 125,048 patients have been confirmed to be infected with SARS-CoV-2 in 117 countries/regions[17]. Therefore, accurate identification and subsequent isolation of NCP patients plays a vital role in blocking the spread of the virus.

Currently, the most commonly used golden standard for NCP diagnosis is a positive result of the real-time RT-PCR assay. However, Ai et al. reported that 3 suspected patients were negative in real-time RT-PCR assay and were later confirmed by metagenomics sequencing[9]. Wang et al. also reported that some patients were negative in the first three tests and turned to positive on the fourth test[18]. In this study, we confirmed 33 patients infected with SARS-CoV-2 from 129 suspected patients. 31(93.94%) of the patients were positive in the first RT-PCR test. Moreover, none of the patients who was negative in our first two tests were later diagnosed as NCP after discharge from hospital. The high positive rate of our assay might be due to the fact that we collected and detected both sputum and pharyngeal swab samples. Furthermore, we detected SARS-CoV-2 with two different kits simultaneously, and only if both results were negative was the patient excluded from SARS-CoV-2 infection. If the results of the two reagents are not consistent, we will use another reagent for re-examination. These measures significantly improved the accuracy of our diagnosis for SARS-CoV-2 infection and NCP.

In our study, 13 RTPs detections were performed in 33 NCP patients, and 2 (6.06%) patients were found to be co-infected with adenovirus or rhinovirus. Both of these two patients didn’t develop into severe conditions, but recovered and were discharged 20 and 24 days after being treated with Coolidge plus Ribavirin. Our findings were consistent with a recent study which reported that none of 99 patients had co-infection of other respiratory viruses[4]. However, Ai et al. reported a high co-infection rate by using mNGS assay which found 5 out of 20 NCP patients co-infected with other viruses[9].

Of the 96 non-NCP patients, 31.25% (30/96) patients were detected at least one respiratory tract pathogen by the SureX 13 respiratory pathogen multiplex kit. The most common pathogen was influenza (including influenza A and B), followed by *Mycoplasma pneumoniae*, and *metapneumovirus*. Our findings were consistent with previous studies that reported the detection rate of pathogens in CAP adults as 30-40%[19]. Respiratory viruses were detected more frequently than bacteria. The most common pathogens in winter were human rhinovirus and influenza virus[19, 20]. In addition, about 70% of the 96 non-NCP patients in this study did not have any specific etiological diagnosis, which may be due to the specimens that we used for the detection being pharyngeal swabs but not alveolar lavage fluid. The positive rate of pharyngeal swab specimen in diagnosing CAP is usually low[20]. The SureX 13 respiratory pathogen multiplex kit used in this study did not include all common CAP pathogens such as *Legionella* and *Chlamydia psittaci*. It was reported that detection of alveolar lavage fluid with mNGS was able to detect more respiratory pathogens in CAP patients ^[21]^. However, due to the difficulty in obtaining such samples and the high cost of the detection, mNGS was not used in this study.

Previous studies reported that secondary bacterial infection commonly occurs in influenza infection and is associated with an increased risk of death[22, 23]. In 2009 pandemic influenza A cases, 18% to 34% of patients admitted to the ICU also had bacterial infection[22]. At present, there are few reports about the incidence of secondary bacterial or fungal infection in NCP patients during treatment. Chen et al. found that none of 99 patients had co-infection of other respiratory viruses, but 4 patients had co-infection of bacteria and fungi. Common pathogens included *A baumannii, K pneumoniae, A flavus, C glabrata*, and *C albicans*. The authors believed that the infection probably occurred secondary to severe ARDS and/or high dose or prolonged use of glucocorticoid therapy[4]. In another study, Ai et al. found 6 cases with bacterial and fungal infection among 20 NCP patients[9]. In our study, we did not find any NCP patients with secondary bacterial or fungal infection during treatment. Although 6 patients’ G test results were above the normal reference value, they were all in severe condition and were given large amounts of albumin and gamma globulin which possibly affected the test results. Moreover, all of the fungal cultures were negative, and the patients did not receive any anti-fungal treatment but still recovered. We suspect that the reason for the low occurrence of secondary infection in our NCP patients is possibly due to the non-invasive regimens, such as maintaining the oxygen saturation through High-flow nasal cannula oxygen therapy (HFNC), but not mechanical ventilation. The patients did not receive peripherally inserted central catheter (PICC), either. Another possible reason is that glucocorticoid was very carefully used to treat the patients even in severe conditions. Our results suggest that the abuse of antimicrobial agents should be avoided in the treatment of patients with SARS-CoV-2.

Earlier studies on NCP patients in Wuhan reported high mortality as 11.1%-14.6%[1, 4]. However, recent studies based on large sample sizes with cases throughout China found a markedly lower case fatality rate (3.92%)[17]. In this study, the mortality of our 85 laboratory-confirmed NCP patients was only 1.18%. This finding is consistent with a most recent study, reporting the mortality of 1.4% of 1,099 cases[8].

In conclusion, in the present study, we found a certain proportion of infection and co-infection of other common pathogens of CAP during the NCP epidemic in Anhui province. This suggested that improvement of accuracy in microbiological laboratory detection is necessary to guide specific and precise treatment for the CAP patients. In this study, no secondary bacterial or fungal infection was detected in NCP patients during the treatment, suggesting that antibiotics should be used cautiously in the treatment of patients with SARS-CoV-2 infection. Please note that this is a single-center study with limited number of patients. The conclusions of this study need to be further verified by expanding the sample size.

## Data Availability

his manuscript has not been published or presented elsewhere in part or in entirety and is not under consideration by another journal.

